# Postpartum anemia and maternal wellbeing: A cohort analysis of the WOMAN-2 trial

**DOI:** 10.1101/2025.11.24.25340880

**Authors:** Judith Lieber, Katharine Ker, Folasade Adenike Bello, Rizwana Chaudhri, Kiran Javaid, Aasia Kayani, Mwansa Ketty Lubeya, Projestine Muganyizi, Oladapo Olayemi, Bellington Vwalika, Monica Arribas, Eni Balogun, Amy Brenner, Amber Geer, Poppy Mallinson, Raoul Mansukhani, Danielle Prowse, Haleema Shakur-Still, Ian Roberts, The WOMAN-2 Trial Collaborators

## Abstract

**Objective:** We examined the association between postpartum anemia and maternal wellbeing in women who gave birth with anemia.

**Methods:** We conducted a cohort analysis using data from the WOMAN-2 trial. Women with moderate or severe anemia who were giving birth vaginally were recruited from hospitals in Nigeria, Pakistan, Tanzania, and Zambia. Our exposure was postpartum anemia (hemoglobin concentration <70, 70-99, or ≥100 g/L (severe, mild/moderate, and no postpartum anemia, respectively)). Our primary outcome was physical capacity (six-minute walk test). Our secondary outcomes were patient-reported vigor, fatigue (general, physical, emotional, mental), overall fatigue, other anemia symptoms, breastfeeding difficulties, expected difficulties with usual activities, breathlessness, illness, and pain. Hemoglobin was measured at 24-hours after birth or discharge and outcomes were measured at discharge or 42 days postpartum, whichever happened first. We assessed the association between postpartum anemia and maternal wellbeing with multivariable regression models.

**Results:** Among 15,068 participants, 11% had severe, 75% had mild/moderate, and 14% had no postpartum anemia. After adjusting for potential confounders, each 10 g/L increase in hemoglobin was associated with 2.99 (95% CI: 2.12 to 3.86) more meters walked in six-minutes. Compared to mild/moderate postpartum anemia, severe anemia was associated with expected difficulties doing usual activities (adjusted odds ratio (aOR)=1.48 (95% CI: 1.14 to 1.91)) and other adverse wellbeing outcomes. No postpartum anemia was associated with less illness (aOR=0.58 (95% CI: 0.35 to 0.96)) and some adverse wellbeing outcomes.

**Conclusion:** Low postpartum hemoglobin was associated with worse wellbeing of new mothers. Our results support recommendations to prevent and treat low postpartum hemoglobin.

## Introduction

Anemia can affect over 50% of postpartum women in low-and-middle-income settings and up to 50% of postpartum women in high-income settings.[1,2] Antepartum anemia and blood loss during childbirth are the main causes of postpartum anemia. The burden of postpartum anaemia is greater in low-and-middle-income settings because antepartum anemia is more common and severe. In South Asia and west and central Africa, a quarter of pregnant women have moderate or severe anemia and another quarter are mildly anemic.[3]

Anemia reduces the oxygen carrying capacity of the blood, which can impair tissue function.[4,5] Postpartum anemia is associated with adverse maternal health and wellbeing outcomes such as fatigue, reduced physical capacity, postpartum depression, and mortality.[6–9] Postpartum anemia may also reduce mothers’ ability to bond with and care for their infant.[10]

Despite its importance in low-and-middle income settings, evidence on the impact of postpartum anemia is largely from small studies in high-income settings.[6–8] We examined the association between postpartum anemia and maternal wellbeing in a large cohort of women who gave birth with moderate or severe anemia in Nigeria, Tanzania, Pakistan, and Zambia.

## Methods

We conducted a cohort analysis using data from the WOMAN-2 trial. Women in active labor were recruited from 34 hospitals in Nigeria, Pakistan, Tanzania, and Zambia. Hemoglobin concentration was measured using a standard point-of-care assessment (HemoCue Hb 201+ System) on women’s arrival at hospital.[11] Women with moderate or severe anemia who were giving birth vaginally were eligible to take part. In line with World Health Organization (WHO) cut-offs, moderate antepartum anemia was defined as a hemoglobin concentration between 70 and 99 g/L and severe antepartum anemia as a hemoglobin concentration below 70 g/L.[12] Women who were under 18 and lacked consent from a guardian, had a known allergy to the trial treatment, had an indication or contraindication to the intervention, or were diagnosed with postpartum hemorrhage before the umbilical cord was clamped were not eligible to take part in the trial. The WOMAN-2 trial protocol and results are published elsewhere.[13,14]

Our exposure was postpartum anemia, defined as postpartum hemoglobin concentration <70, 70-99, or ≥100 g/L (severe, mild/moderate and no postpartum anemia, respectively)(20). Hemoglobin was measured with the HemoCue 24-hours after birth or at discharge, whichever happened first.

Our primary outcome was physical capacity, which we assessed with the six-minute walk test. This is an objective and validated measure of physical capacity that jointly assesses the cardiovascular, neuromuscular, and other physiological systems.[15] Our secondary outcomes were patient-reported vigor, fatigue (general, physical, emotional, mental), overall fatigue, other anemia symptoms, breastfeeding difficulties, expected difficulties with usual activities, post-walk breathlessness, illness, and pain (table S1). The outcomes were measured at discharge or day 42, whichever happened first. Physical capacity and vigor represent ‘good’ maternal wellbeing, while the remaining outcomes represent ‘poor’ maternal wellbeing.

We measured vigor and fatigue (general, physical, emotional, mental) with subscales of the Multi-dimensional Fatigue Symptom Inventory: Short Form (MFSI-SF). This was adapted for postpartum women.[16] The vigor subscale assesses positive feelings (e.g., ‘Taking into account everything since giving birth until now, I feel happy’) (table S1). The general fatigue subscale assesses tiredness (e.g., ‘Since giving birth until now, I feel physically and mentally tired). The physical fatigue subscale assesses body and muscle weakness (e.g., ‘Since giving birth until now, my legs feel weak’). The mental fatigue subscale assesses cognition (e.g., ‘Taking into account everything since giving birth until now, my mind is wandering around). The emotional fatigue subscale assesses negative emotions (e.g., ‘Taking into account everything since giving birth until now, I feel depressed’). We calculated each score by summing the subscale’s six Likert-style questions (range 0 to 24).[16] We measured overall fatigue by subtracting the vigor score from the sum of the general, physical, mental, and emotional fatigue scores (range –24 to 94).[16] We dichotomized this score (≥1 representing overall fatigue).

The remaining patient-reported outcomes were developed for, and validated in, women with postpartum anemia.[16] We measured other anemia symptoms by summing the relevant seven Likert-style questions (range 0 to 28) (table S1). For breastfeeding difficulties, expected difficulties with usual activities, post-walk breathlessness, illness, and pain, women were defined as having the outcome if they responded ‘quite a bit’ or ‘extremely’ to the relevant Likert-style question (table S1).

Women who died were categorised as having the ‘worst’ observation for each outcome. Women who had not yet breastfed were categorised as having ‘extreme’ difficulties with breastfeeding. We excluded women who had a baby in the neonatal intensive care unit or perinatal death from analyses of breastfeeding difficulties.

We developed a directed acyclic graph to identify potential confounders (figure S1). We examined the association between postpartum anemia and physical capacity with linear regression, adjusting for potential confounders. These include sociodemographics, pregnancy history, maternal complications, birth and baby characteristics (table S2 for a full list of covariates). We also adjusted for time at outcome assessment. We did not adjust for surgical and non-surgical interventions for bleeding (figure S1) because they were closely correlated with clinical diagnosis of postpartum hemorrhage. We used negative binomial regression to model the count outcomes (fatigue subscales and other anemia symptoms) and logistic regression to model the binary outcomes. We included a random-effect at the hospital-level to account for correlations between women attending the same hospital.

In a sensitivity analysis, we modelled the count measures as binary outcomes using logistic regression. Women were defined as having vigor, fatigue (general, physical, emotional, mental), or other anemia symptoms if they responded ‘quite a bit’ or ‘extremely’ to any of the relevant Likert-style questions (table S1). In a further sensitivity analysis, we assessed whether postpartum anemia was associated with maternal wellbeing via time at discharge (figure S1) by removing time at outcome assessment from our multivariable models.

We used restricted cubic splines (four knots) to explore the shape of the relationship between postpartum hemoglobin concentration and each outcome. We present this as adjusted predictions of physical capacity or score measures (or risks for the binary outcomes) at different concentrations of postpartum hemoglobin. We did a complete-case analysis as there were little missing data.

We used STATA 18 and R (version 4.2.0) for our analyses.

## Ethics

The WOMAN-2 protocol was approved by the National Health Research Ethics Committee of Nigeria (NHREC/01/01/2007-29/09/2019); the National Bioethics Committee of Pakistan (NBC-340); the National Institute of Medical Research of Tanzania (NIMR/HQ/R.8a/Vol.IX/3767); the University of Zambia Biomedical Research Ethics Committee (REF 001-04-19); and the London School of Hygiene and Tropical Medicine’s Ethics Committee (15194). A full description of consent processes is published elsewhere.[13,14]

## Results

A total of 15,068 women took part in the WOMAN-2 trial between August 2019 and September 2023. There were missing data for postpartum anemia (N=205), time to outcome assessment (N=258), six-minute walk test and post-walk breathlessness (N=580, respectively), breastfeeding difficulties (N=287), expected difficulties with usual activities (N=245), illness and pain (N=240, respectively), labor duration (N=85), small vulnerable newborn (N=32), estimated blood loss (N=10), perinatal death (N=6), maternal death, clinical diagnosis of postpartum hemorrhage, and parity (N=2, respectively). Table S3 shows the characteristics of women who are missing postpartum anemia data. There were complete data for the other variables of interest.

In the cohort, 11% of women had severe postpartum anemia, 75% had mild/moderate postpartum anemia, and 14% were not anemic after birth, 7% were diagnosed with a postpartum hemorrhage, and 25% received a blood transfusion (table 1). Mean postpartum hemoglobin concentration was 85.8 (standard deviation (SD) 13.6)). Eighteen women died. Table S3 shows the participants’ characteristics by postpartum anemia status. Severe pre-birth anemia was least common in women who were not anemic after birth (5% compared to 11% and 46% in women with mild/moderate and severe postpartum anemia, respectively). Blood transfusion was most common in these women (39% compared to 30% and 22% in women with severe and mild/moderate postpartum anemia, respectively).

**Table 1.** Participants’ characteristics.

|  | **N (%)** | **Total N** |
| --- | --- | --- |
|  | **N=15,068** | **15,068** |
| Postpartum hemoglobin (g/L) (mean, SD) | 85.78 (13.61) | 14,863 |
| Postpartum anemia |  | 14,863 |
| No anemia ($\geq 100$ g/L) | 2,097 (14.11%) | |
| Mild-Moderate (70-99 g/L) | 11,107 (74.73%) |  |
| Severe ($< 70$ g/L) | 1,659 (11.16%) | |
| Maternal age (years) (mean, SD) | 27.20 (5.61) | 15,068 |
| Maternal age (years) |  | 15,068 |
| $< 20$ | 895 (5.94%) | |
| 20-29 | 8,937 (59.31%) |  |
| 30-39 | 4,830 (32.05%) |  |
| 40+ | 406 (2.69%) |  |
| Country |  | 15,068 |
| Nigeria | 1,326 (8.80%) |  |
| Pakistan | 11,025 (73.17%) |  |
| Tanzania | 2,029 (13.47%) |  |
| Zambia | 688 (4.57%) |  |
| Hemoglobin at baseline (g/L) (mean, SD) | 82.74 (11.84) | 15,068 |
| Anemia severity at baseline |  | 15,068 |
| Moderate | 12,989 (86.20%) |  |
| Severe | 2,079 (13.80%) |  |
| Multiple gestation | 583 (3.87%) | 15,068 |
| Gestational age (weeks) (mean, SD) | 37.39 (2.68) | 15,068 |
| Primiparity | 5,031 (33.39%) | 15,066 |
| Long-term infection | 552 (3.66%) | 15,068 |
| Placental abnormalities | 469 (3.11%) | 15,068 |
| Hypertensive disorder | 1,146 (7.61%) | 15,068 |
| Small vulnerable newborn | 5,205 (34.62%) | 15,036 |
| Labor induction | 1,831 (12.15%) | 15,068 |
| Labor augmentation | 4,493 (29.82%) | 15,068 |
| Instrumental delivery | 422 (2.80%) | 15,068 |
| Episiotomy | 4,355 (28.90%) | 15,068 |
| Pharmacological pain relief | 1,810 (12.01%) | 15,068 |
| Duration of labor (hours) (median, IQR) | 9.18 (6.42-13.75) | 14,983 |
| Injury to the birth canal | 2,190 (14.53%) | 15,068 |
| Estimated blood loss (ml) (mean, SD) | 310.31 (192.72) | 15,058 |
| Clinically diagnosed PPH | 1,034 (6.86%) | 15,066 |
| Intervention for PPH | 485 (3.22%) | 15,066 |
| Postpartum blood transfusion | 3,822 (25.37%) | 15,068 |
| Postpartum intravenous fluids | 4,764 (31.62%) | 15,068 |
| Baby admitted to NICU | 193 (1.28%) | 15,068 |
| Perinatal death before discharge | 1,172 (7.78%) | 15,062 |
| Small vulnerable newborn | 5,205 (34.62%) | 15,036 |
| Maternal death | 18 (0.12%) | 15,066 |
| Time to outcome assessment (hours)<br>(median, IQR) | 12.34 (6.19-23.92) | 15,019 |
| SD – standard deviation; IQR – interquartile range; PPH – postpartum hemorrhage;<br>NICU – neonatal intensive care unit. |  |  |

Hemoglobin concentration was mostly measured on the first postpartum day (median 12.6 hours (interquartile range (IQR) 6.5 to 23.6)), as were the wellbeing outcomes (median 12.3 hours (IQR 6.2 to 23.9)). On average, women walked 212.2 meters during the six-minute walk test (SD 92.8) (table S4). The most common patient-reported outcomes were vigor (75.7%), overall fatigue (26.2%), and difficulties breastfeeding (9.9%), and least common were illness (1.4%), pain (1.3%), and post-walk breathlessness (0.9%) (table S4).

Table 2 shows the adjusted associations between postpartum anemia and wellbeing outcomes (table S5 for unadjusted results). Compared to mild/moderate postpartum anemia, severe anemia was associated with lower physical capacity (adjusted β-coefficient –3.56 meters (95% CI: –7.17 to 0.04)) and no postpartum anemia was associated with greater physical capacity (6.37 meters (95% CI: 3.28 to 9.46)). We modelled hemoglobin concentration as a continuous exposure for physical capacity and post-walk breathlessness because the flexible spline models show approximately linear relationships (figure 1). This was supported by lower AIC and BIC values of the linear models, which represents better model fit. After adjusting for potential confounders, each 10 g/L increase in postpartum hemoglobin was associated with 2.99 (95% CI: 2.12 to 3.86) more meters walked within six-minutes and 40% lower odds of post-walk breathlessness (adjusted odds ratio (aOR) = 0.60 (95% CI: 0.49 to 0.74)) (not shown). Women who did not participate in the six-minute walk had worse postpartum anemia (table S6).

**Figure 1.**
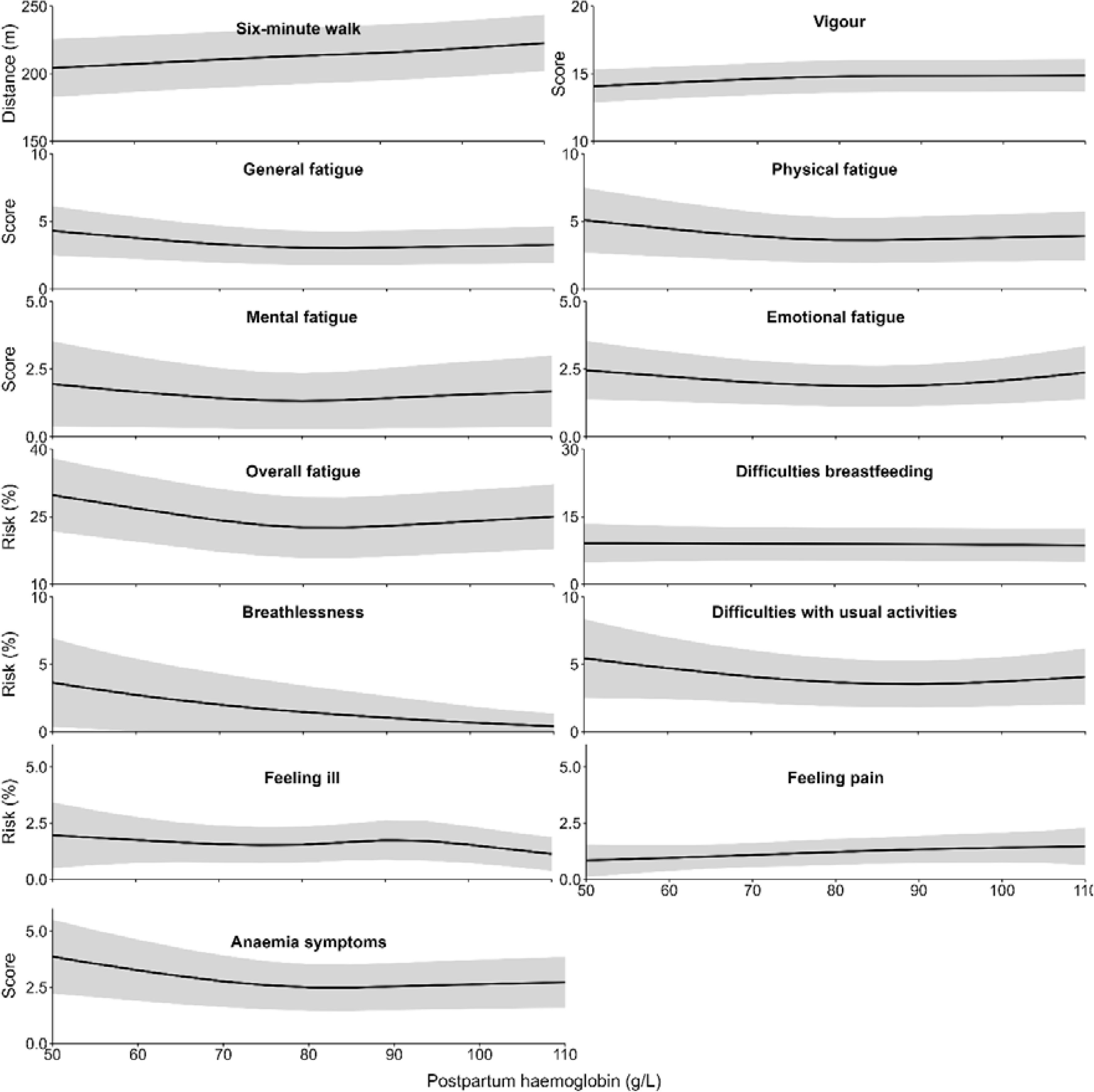
Associations between postpartum hemoglobin concentration (g/L) and physical capacity (six-minute walk), vigor, fatigue (general, physical, emotional, mental), overall fatigue, difficulties breastfeeding, post-walk breathlessness, expected difficulties with usual activities, illness, pain, and anemia symptoms. Graphs of model-based predictions from linear regression (physical capacity), negative binomial regression (vigor, general, physical, emotional, and mental fatigue, other anemia symptoms), and logistic regression (overall fatigue, difficulties breastfeeding, post-walk breathlessness, expected difficulties with usual activities, illness, and pain) models treating the exposure (postpartum hemoglobin concentration (g/L)) as a restricted cubic spline with four knots. Models were adjusted for time to outcome assessment, pre-birth anemia severity, age, gestational age, multiple gestation, primiparity, placental abnormalities, long-term infection, hypertensive disorder, labor induction, labor augmentation, pain relief, instrumental birth, episiotomy, trauma to birth canal, labor duration, estimated total blood loss (quadratic term), clinical diagnosis of postpartum hemorrhage, postpartum intravenous fluids, postpartum blood transfusion, small vulnerable newborn, and perinatal death before discharge.

**Table 2.** Adjusted association between postpartum anemia and physical capacity, vigor, fatigue (general, physical, emotional, mental), other anemia symptoms, overall fatigue, difficulties breastfeeding, expected difficulties with usual activities, post-walk breathlessness, illness, and pain.

|  | Postpartum anemia |  |  |
| --- | --- | --- | --- |
|  | Severe | Mild/<br>Moderate | None |
|  | <u>β-coefficient (95% CI)</u> |  |  |
| Physical capacity (six-minute walk) | -3.56 (-7.17 to 0.04) | 0.00 | 6.37 (3.28 to 9.46) |
|  | <u>Incident rate ratio (95% CI)</u> |  |  |
| Vigor | 0.99 (0.97 to 1.01) | 1.00 | 1.00 (0.99 to 1.02) |
| General fatigue | 1.16 (1.09 to 1.23) | 1.00 | 1.06 (1.00 to 1.11) |
| Physical fatigue | 1.16 (1.09 to 1.23) | 1.00 | 1.08 (1.03 to 1.13) |
| Emotional fatigue | 1.10 (0.98 to 1.24) | 1.00 | 1.21 (1.09 to 1.34) |
| Mental fatigue | 1.18 (1.03 to 1.36) | 1.00 | 1.18 (1.05 to 1.36) |
| Other anemia symptoms | 1.21 (1.13 to 1.29) | 1.00 | 1.06 (1.01 to 1.12) |
|  | <u>Odds ratio (95% CI)</u> |  |  |
| Overall fatigue | 1.22 (1.04 to 1.42) | 1.00 | 1.17 (1.02 to 1.34) |
| Difficulties with breastfeeding | 0.95 (0.75 to 1.21) | 1.00 | 1.00 (0.83 to 1.22) |
| Expected difficulties with usual activities | 1.48 (1.14 to 1.91) | 1.00 | 0.97 (0.73 to 1.27) |
| Post-walk breathlessness |  | 1.00 |  |
| Illness | 0.98 (0.63 to 1.53) | 1.00 | 0.58 (0.35 to 0.96) |
| Pain | 0.75 (0.45 to 1.26) | 1.00 | 1.11 (0.70 to 1.76) |
| All models included a random effect for hospital and adjust for time to outcome assessment, pre-birth anemia severity, age, gestational age, multiple gestation, primiparity, placental abnormalities, long-term infection, hypertensive disorder, labor induction, labor augmentation, pain relief, instrumental birth, episiotomy, trauma to birth canal, labor duration, estimated total blood loss (quadratic term), clinical diagnosis of postpartum hemorrhage, postpartum intravenous fluids, postpartum blood transfusion, small vulnerable newborn, and perinatal death before discharge. CI – confidence interval. |  |  |  |

Compared to mild/moderate postpartum anemia, severe anemia was associated with expected difficulties with usual activities (aOR=1.48 (95% CI: 1.14 to 1.91), overall fatigue (aOR=1.22 (95% CI: 1.04 to 1.42), other anemia symptoms (adjusted incident rate ratio (aIRR)=1.21 (95% CI: 1.13 to 1.29)), mental fatigue (aIRR=1.18 (95% CI: 1.03 to 1.36)), general fatigue (aIRR=1.16 (95% CI: 1.09 to 1.23)), physical fatigue (aIRR=1.16 (95% CI: 1.09 to 1.23)), and emotional fatigue (aIRR=1.10 (95% CI: 0.98 to 1.24)) (table 2). Compared to mild/moderate postpartum anemia, no anemia was associated with less reported illness (aOR=0.58 (95% CI: 0.35 to 0.96)) and greater emotional fatigue (aIRR=1.21 (95% CI: 1.09 to 1.34)), mental fatigue (aIRR=1.18 (95% CI: 1.05 to 1.36)), physical fatigue (aIRR=1.08 (95% CI: 1.03 to 1.13)), other anemia symptoms (aIRR=1.06 (95% CI: 1.01 to 1.12)), general fatigue (aIRR=1.06 (95% CI: 1.00 to 1.11)), and overall fatigue (aOR=1.17 (95% CI: 1.02 to 1.34)). These non-linear relationships are shown in the cubic spline models (figure 1). We found no evidence that postpartum anemia was associated with vigor, breastfeeding difficulties, or pain (table 2).

We observed similar results when we modelled the score measures as binary outcomes using logistic regression (tables S7-8) and when we removed time at outcome assessment from our multivariable models (table S9).

## Discussion

Lower postpartum hemoglobin was associated with worse physical capacity of new mothers. Women with severe postpartum anemia reported worse wellbeing outcomes compared to women with mild/moderate postpartum anemia, while women without postpartum anemia reported mixed outcomes.

We explored the impact of postpartum anaemia on maternal wellbeing in a large cohort of women giving birth with anaemia in Nigeria, Tanzania, Pakistan, and Zambia. We were able to examine the impact of severe postpartum anemia on maternal wellbeing due to its high incidence in the study population. We assessed a range of maternal wellbeing outcomes that women highlighted as important during the postpartum period.[16] These measures were developed for, and validated in, women with postpartum anemia.[16] We measured postpartum hemoglobin with the HemoCue Hb 201+. While this has high accuracy compared with laboratory measurements, some measurement error is expected.[17] Women who did not take part in the six-minute walk had worse postpartum anemia, so we may have underestimated its effect on physical capacity. The patient-reported outcomes were measured at discharge, which may have led to measurement error if women hurried the questionnaire to return home. We explored linear and non-linear relationships with spline models and modelled them appropriately for each outcome. While there are no established thresholds for postpartum anemia, we used British Society of Hematology cut-offs that broadly match the non-linear relationships shown in the spline models.[18,19] We adjusted for a range of potential confounders, including pre-birth anemia severity and postpartum bleeding. However, residual confounding could still bias our results. We undertook multiple sensitivity analyses. These did not impact our findings.

Postpartum hemoglobin was associated with increased physical capacity and reduced breathlessness. This is in line with randomized trials that show increasing hemoglobin improves physical capacity.[7,20] Lower hemoglobin may result in less oxygen delivery to the tissues, which would impact cellular energy production.[4,5] Cardiac and skeletal muscles have high energy demands.

Severe postpartum anemia was associated with a range of adverse maternal wellbeing outcomes when compared to mild/moderate anemia. This may result from compensatory mechanisms that maintain an adequate oxygen supply to the tissues in mild/moderate anemia but fail at lower hemoglobin concentrations.[21] While (to our knowledge) this is the first study to assess the impact of severe anemia in the postpartum period, there is extensive evidence of its adverse impact on pregnant women.[9,22] Severe postpartum anemia is common in women who give birth with anemia (over 10% of the study cohort) and/or bleed heavily during childbirth (e.g., caesarean births).[23]

Although the study was restricted to women with moderate or severe anemia before birth, 14% of women were not anemic after birth. These women had the highest hemoglobin before birth, which may have been raised above 100 g/L by hemoconcentration and/or blood transfusion. It is also possible their true pre-birth hemoglobin was ≥100 g/L. Compared to mild/moderate postpartum anemia, no postpartum anemia was associated with less reported illness but greater emotional and mental fatigue. This conflicts with randomized trials that show treating anemia reduces postpartum depression and fatigue.[6,8] Our results could be confounded by pre-birth blood transfusion and/or peripartum iron supplementation, which we were unable to adjust for. Differential misclassification may also have occurred. Women who had mild/moderate postpartum anemia were more anemic before birth, so could have normalized fatigue and reported their symptoms less. We recommend interpreting these U-shaped relationships with caution.

We found no evidence that postpartum anemia was associated with vigor, breastfeeding difficulties, or pain. These outcomes have diverse causes that may have impacted women’s wellbeing more than their anemia status. For example, breastfeeding difficulties can occur because of the baby’s status as well as the mother’s.

Future studies could examine the impact of postpartum anemia on maternal wellbeing in women who are not anemic before birth. Hemoglobin was measured around 12 hours after birth. While this is in line with guidelines for women with antepartum anemia, hemoglobin is highly variable in this period.[18] Maternal wellbeing was measured before discharge from hospital. Future studies could examine the effect of postpartum anemia on long-term outcomes like postpartum depression and mother-infant bonding, measuring hemoglobin after 48 hours postpartum.

## Conclusion

Low postpartum hemoglobin was associated with adverse wellbeing in new mothers. While we cannot infer causation from this observational study, this in line with randomized controlled trials that show treating anemia improves a range of wellbeing outcomes.[7,8,20] These results support recommendations to prevent and correct low postpartum hemoglobin.[18,24]

## Author contributions

JL did the conceptualization, methodology, formal statistical analysis, visualization, and writing of the original draft. IR did the conceptualization, methodology, supervision, funding acquisition, writing of the original draft and draft revisions. HSS did the funding acquisition and review and editing of the manuscript. KK did the writing of the original draft and draft revisions. FAB, RC, KJ, AK, MKL, PM, OO, and BV did the data collection, data interpretation, and review and editing of the manuscript. EB and MA did the project administration and review and editing of the manuscript. AG and DP did the data curation and review and editing of the manuscript. PM did the methodology and review and editing of the manuscript. AB and RM did the review and editing of the manuscript. JL, AG, DP, and RM, have accessed and verified the data. All authors are responsible for the decision to submit the manuscript.

## AI disclosure

ChatGPT (https://chat.openai.com/) was used to reword short phrases.

## Data Availability

Individual deidentified WOMAN-2 trial patient data, including a data dictionary, will be made available via our data sharing portal, The Free Bank of Injury and Emergency Research Data (freebird.lshtm.ac.uk) website.

https://www.freebird.lshtm.ac.uk

## Acknowledgements

We would like to thank all the study sites and staff who made the WOMAN-2 trial happen and most importantly, the participating women, without whom the study would not have been possible.

## References

[1] Mzembe G, Moya E, Mwangi MN, Ataide R, Harding R, Kaunda J, et al. Postpartum maternal and infant haematological effects of second-trimester ferric carboxymaltose versus standard-of-care oral iron in Malawi: longitudinal follow-up of a randomised controlled trial. Lancet Glob Heal 2024;12:e2049–58. 10.1016/S2214-109X(24)00380-2.

[2] Churchill D, Ali H, Moussa M, Donohue C, Pavord S, Robinson SE, et al. Maternal iron deficiency anaemia in pregnancy: Lessons from a national audit. Br J Haematol 2022;199:277–84. 10.1111/bjh.18391.

[3] Stevens GA, Paciorek CJ, Flores-Urrutia MC, Borghi E, Namaste S, Wirth JP, et al. National, regional, and global estimates of anaemia by severity in women and children for 2000–19: a pooled analysis of population-representative data. Lancet Glob Heal 2022;10:e627–39. 10.1016/S2214-109X(22)00084-5.

[4] Weiskopf RB, Feiner J, Hopf HW, Viele MK, Watson JJ, Kramer JH, et al. Oxygen reverses deficits of cognitive function and memory and increased heart rate induced by acute severe isovolemic anemia. Anesthesiology 2002;96:871–7. 10.1097/00000542-200204000-00014.

[5] Toy P, Feiner J, Viele MK, Watson J, Yeap H, Weiskopf RB. Fatigue during acute isovolemic anemia in healthy, resting humans. Transfusion 2000;40:457–60. 10.1046/j.1537-2995.2000.40040457.x.

[6] Moya E, Phiri N, Choko AT, Mwangi MN, Phiri KS. Effect of postpartum anaemia on maternal health 17 related quality of life[: a systematic review and meta 17 analysis. BMC Public Health 2022:1–10. 10.1186/s12889-022-12710-2.

[7] Prick BW, Jansen AJG, Steegers EAP, Hop WCJ, Essink-Bot ML, Uyl-De Groot CA, et al. Transfusion policy after severe postpartum haemorrhage: A randomised non-inferiority trial. BJOG An Int J Obstet Gynaecol 2014;121:1005–14. 10.1111/1471-0528.12531.

[8] Jensen MCH, Holm C, Jørgensen KJ SJ. Treatment for women with postpartum iron deficiency anaemia. Cochrane Database Syst Rev 2024. 10.1002/14651858.cd004222.

[9] Daru J, Zamora J, Fernández-Félix BM, Vogel J, Oladapo OT, Morisaki N, et al. Risk of maternal mortality in women with severe anaemia during pregnancy and post partum: a multilevel analysis. Lancet Glob Heal 2018;6:e548–54. 10.1016/S2214-109X(18)30078-0.

[10] Murray-Kolb LE, Beard JL. Iron deficiency and child and maternal health. Am J Clin Nutr 2009;89:946S–950S. 10.3945/ajcn.2008.26692D.

[11] HemoCue. HemoCue Hb 201+ system. n.d. https://www.hemocue.com/en/solutions/hematology/hemocue-hb-201plus-system (accessed January 28, 2025).

[12] World Health Organization. Guideline on haemoglobin cutoffs to define anaemia in individuals and populations. Geneva: 2024.

[13] WOMAN-2 Trial Collaborators. The effect of tranexamic acid on postpartum bleeding in women with moderate and severe anaemia (WOMAN-2): an international, randomised, double blind, placebo-controlled trial. Lancet 2024;404:1645–56. 10.1016/S0140-6736(24)01749-5.

[14] Ker K, Roberts I, Chaudhri R, Fawole B, Beaumont D, Balogun E, et al. Tranexamic acid for the prevention of postpartum bleeding in women with anaemia: Study protocol for an international, randomised, double-blind, placebo-controlled trial. Trials 2018;19:1–19. 10.1186/s13063-018-3081-x.

[15] American Thoracic Society. ATS Statement□ Guidelines for the Six-Minute Walk Test. Am J Respir Crit Care Med 2002;166:111–7. 10.1164/rccm.166/1/111.

[16] Miller L, Chaudhri S, Beaumont D, Kayani A, Javid K, Chaudhri R, et al. Development of a patient reported outcome questionnaire to measure the impact of postpartum blood loss in women with moderate and severe anaemia: A study using a multi-faceted approach. Wellcome Open Res 2019;4. 10.12688/wellcomeopenres.15245.1.

[17] Bell S, Sweeting M, Ramond A, Chung R, Kaptoge S, Walker M, et al. Comparison of four methods to measure haemoglobin concentrations in whole blood donors (COMPARE): A diagnostic accuracy study. Transfus Med 2021;31:94–103. 10.1111/tme.12750.

[18] Pavord S, Daru J, Prasannan N, Robinson S, Stanworth S, Girling J. UK guidelines on the management of iron deficiency in pregnancy. Br J Haematol 2020;188:819–30. 10.1111/bjh.16221.

[19] Yourkavitch J, Obara H, Usmanova G, Semrau KEA, Moller AB, Garcia-Casal MN, et al. A rapid landscape review of postpartum anaemia measurement: challenges and opportunities. BMC Public Health 2023;23:1–11. 10.1186/s12889-023-16383-3.

[20] Pasricha SR, Low M, Thompson J, Farrell A, De-Regil LM. Iron supplementation benefits physical performance in women of reproductive age: A systematic review and meta-analysis. J Nutr 2014;144:906–14. 10.3945/jn.113.189589.

[21] Weiskopf RB, Viele MK, Feiner J, Kelley S, Lieberman J, Noorani M, et al. Human cardiovascular and metabolic response to acute, severe isovolemic anemia. Jama 1998;279:217–21. 10.1001/jama.279.3.217.

[22] Young MF, Oaks BM, Tandon S, Martorell R, Dewey KG, Wendt AS. Maternal hemoglobin concentrations across pregnancy and maternal and child health: a systematic review and meta-analysis. Ann N Y Acad Sci 2019;1450:47–68. 10.1111/nyas.14093.

[23] Butwick AJ, Walsh EM, Kuzniewicz M, Li SX, Escobar GJ. Patterns and Predictors of Severe Postpartum Anemia after Cesarean Section. Transfusion 2017;176:100–106. 10.1177/0022146515594631.Marriage.

[24] Muñoz M, Stensballe J, Ducloy-Bouthors AS, Bonnet MP, De Robertis E, Fornet I, et al. Patient blood management in obstetrics: Prevention and treatment of postpartum haemorrhage. A NATA consensus statement: A multidisciplinary consensus statement. Blood Transfus 2019;17:112–36. 10.2450/2019.0245-18.

