## Supplementary materials for "Postpartum anemia and maternal wellbeing: A cohort analysis of the WOMAN-2 trial"

Table S1. Patient-reported outcome questionnaire and corresponding maternal wellbeing outcomes

| **Questionnaire item** | **Response categories** | **Study outcome** |
| --- | --- | --- |
| I1. I have pain right now | Not at all (0) / A little (1) / Moderately (2) / Quite a bit (3) / Extremely (4) | Pain |
| I2. I feel ill right now |  | Illness |
| J1. Since giving birth until now, I have trouble remembering things |  | Mental fatigue |
| J2. Since giving birth until now, my muscles ache all over my body (muscles ache) |  | Physical fatigue |
| J3. Taking into account everything since giving birth until now, I feel upset |  | Emotional fatigue |
| J4. Since giving birth until now, my legs feel weak |  | Physical fatigue |
| J5. Taking into account everything since giving birth until now, I feel happy (cheerful) |  | Vigor |
| J6. Since giving birth until now, my head feels heavy |  | Physical fatigue |
| J7. Taking into account everything since giving birth until now, I feel happy to do things with energy (feel lively) |  | Vigor |
| J8. Taking into account everything since giving birth until now, I feel nervous or uneasy |  | Emotional fatigue |
| J9. Taking into account everything since giving birth until now, I feel relaxed |  | Vigor |
| J10. Since giving birth until now, I feel too tired to continue (I feel pooped) |  | General fatigue |
| J11. Since giving birth until now, I am confused or I have difficulty understanding things |  | Mental fatigue |
| J12. Since giving birth until now, I am drained of energy (I am worn out) |  | General fatigue |
| J13. Taking into account everything since giving birth until now, I feel sad |  | Emotional fatigue |
| J14. Since giving birth until now, I feel physically and mentally tired (fatigued) |  | General fatigue |
| J15. Taking into account everything since giving birth until now, my mind is wandering around (I have trouble paying attention) |  | Mental fatigue |
| J16. Since giving birth until now, my arms feel weak |  | Physical fatigue |
| J17. Since giving birth until now, I feel physically slow (sluggish) |  | General fatigue |
| J18. Since giving birth until now, I feel in a bad state (I feel run down) |  | General fatigue |
| J19. Since giving birth until now, I ache all over |  | Physical fatigue |
| J20. Taking into account everything since giving birth until now, I have trouble focusing on things (I am unable to concentrate) |  | Mental fatigue |
| J21. Taking into account everything since giving birth until now, I feel depressed |  | Emotional fatigue |
| J22. Taking into account everything since giving birth until now, I feel refreshed or fresh |  | Vigor |
| J23. Taking into account everything since giving birth until now, I feel tense |  | Emotional fatigue |
| J24. Since giving birth until now, I feel energetic or have energy to do things |  | Vigor |
| J25. Since giving birth until now, I make more mistakes than usual |  | Mental fatigue |
| J26. Since giving birth until now, my body feels heavy all over |  | Physical fatigue |
| J27. Since giving birth until now, I am forgetful |  | Mental fatigue |
| J28. Since giving birth until now, I feel physically tired |  | General fatigue |
| J29. Taking into account everything since giving birth until now, my mind is at peace (I feel calm) |  | Vigor |
| J30. Taking into account everything since giving birth until now, I am very worried (distressed) |  | Emotional fatigue |
| K1. Since giving birth until now, I have felt dizzy |  | Other anemia symptoms |
| K2. Since giving birth until now, I have had a headache |  |  |
| K3. Since giving birth until now, I have felt my heart beating very fast or strangely |  |  |
| K4. Since giving birth until now, I have felt sleepy |  |  |
| K5. Since giving birth until now, I have felt numbness in my hands and feet |  |  |
| K6. Since giving birth until now, my hands have felt shaky |  |  |
| K7. Since giving birth I have had difficulty in breathing |  |  |
| K8a. Have you breastfed your baby since giving birth? | Yes / No / N/a | Breastfeeding difficulties |
| K8b. I have had difficulty breastfeeding my baby | Not at all (0) / A little (1) / Moderately (2) / Quite a bit (3) / Extremely (4) |  |
| K9. When I go home, I will have difficulty doing my usual activities |  | Expected difficulties with usual activities |
| [Please ask the participant immediately after the walk test]  M6. I have difficulty in breathing right now |  | Post-walk breathlessness |

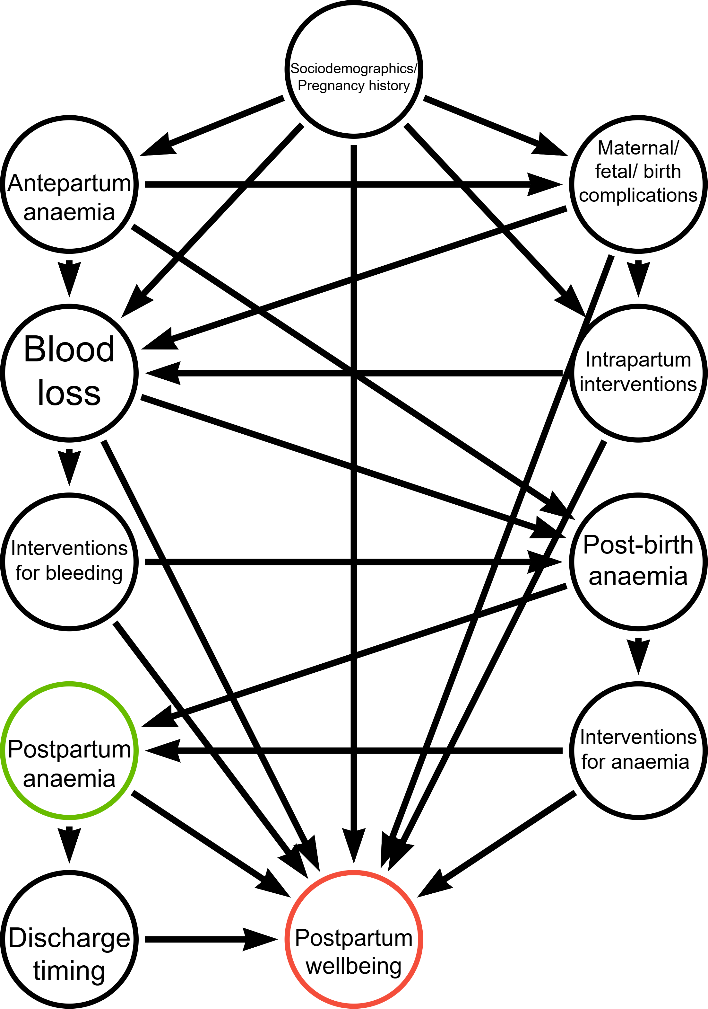
Figure S1. Directed acyclic graph showing the potential causal pathways between postpartum anemia and postpartum wellbeing

The exposure is shown in green and the outcome in red. Sociodemographics/pregnancy history includes maternal age and parity. Maternal/birth/fetal complications include multiple gestation, placental abnormalities, hypertensive disorders, long-term infection, trauma to the birth canal, long labor, small vulnerable newborn, and perinatal death. Intrapartum interventions include labor induction, labor augmentation, pain relief, instrumental delivery, and episiotomy. Bleeding interventions include intravenous fluids, blood transfusion, surgical (e.g., arterial ligation, hysterectomy, laparotomy) and non-surgical treatments (e.g., uterine tamponade, bimanual compression, non-pneumatic anti-shock garment). Interventions for anemia include blood transfusion and iron supplementation.

Table S2. Description of covariates included in the regression models

| **Covariate** | **Definition** | **Data source** |
| --- | --- | --- |
| Time to outcome assessment | Time between birth and measurement of study outcomes (hours) | Measured by research team |
| Pre-birth anemia severity | Maternal hemoglobin concentration before birth: <70 g/dL (severe) or 70-99 g/dL (moderate) anemia |  |
| Maternal age | Maternal age at delivery (years) | Self-reported by mother or documented from medical records |
| Gestational age | Total weeks gestation at birth (weeks) |  |
| Multiple gestation | Mother carrying two or more fetuses in this pregnancy |  |
| Primiparity | First pregnancy carried to viable gestational age |  |
| Placental abnormalities | Placental abruption, previa, accreta, increta, and/or percreta |  |
| Long-term infection | HIV, syphilis, hepatitis, malaria, TB, and/or other infectious disorder |  |
| Hypertensive disorder | Pre-existing hypertension, gestational hypertension, eclampsia, or pre-eclampsia |  |
| Labor induction | Receipt of any method of labor induction (mechanical and/or pharmacological) |  |
| Labor augmentation | Receipt of any method of labor augmentation |  |
| Pain relief | Receipt of any method of pharmacological pain relief during birth |  |
| Instrumental delivery | Forceps, ventouse, or other instrument to support delivery |  |
| Episiotomy | Any cut made to the perineum during birth |  |
| Trauma to the birth canal | Any cut, fracture, or other injury sustained to the cervix, vagina or perineum by the baby during birth |  |
| Estimated blood loss | Estimate of total blood loss (ml) at 24 hours after trial treatment, death or discharge from hospital, whichever occurs first |  |
| Clinically diagnosed postpartum hemorrhage | A diagnosis of postpartum hemorrhage made by the treating clinician. This could be an estimated blood loss of more than 500 mL or any blood loss sufficient to compromise hemodynamic stability within 24 hours of delivery. Hemodynamic instability was based on signs such as tachycardia, reduced urine output, or low systolic blood pressure |  |
| Intravenous fluids | Any intravenous fluids given after umbilical cord clamping/cutting |  |
| Blood transfusion | Any blood transfusion given after umbilical cord clamping/cutting |  |
| Perinatal death | Any stillbirth or neonatal death before discharge |  |
| Total duration of labor | Time between onset of painful contractions and delivery (hours) | Onset time self-reported by mother. Delivery time measured by research team |
| Small vulnerable newborn | Any preterm (<37 weeks) and/or small-for-gestational-age and/or low birthweight (<2,500 g) baby.[25] | Measured by research team or documented from medical records |

Table S3. Participants’ characteristics, by postpartum anemia status

|  | **N (%)** | | | | |
| --- | --- | --- | --- | --- | --- |
|  | **Postpartum anemia** | | | | |
|  | **Severe** | **Mild-Moderate** | **No anemia** | **Missing data** | **Total** |
|  | **N=1,659** | **N=11,107** | **N=2,097** | **N=205** | **N=15,068** |
| Postpartum hemoglobin (g/L) (mean, SD) | 61.87 (7.01) | 85.33 (7.80) | 107.08 (6.71) | - | 85.78 (13.61) |
| Maternal age (years) | 27.13 (5.80) | 27.12 (5.58) | 27.70 (5.60) | 26.60 (5.55) | 27.20 (5.61) |
| Maternal age (years) |  |  |  |  |  |
| <20 | 113 (6.81%) | 659 (5.93%) | 113 (5.39%) | 10 (4.88%) | 895 (5.94%) |
| 20-29 | 961 (57.93%) | 6,662 (59.98%) | 1,180 (56.27%) | 134 (65.37%) | 8,937 (59.31%) |
| 30-39 | 534 (32.19%) | 3,497 (31.48%) | 744 (35.48%) | 55 (26.83%) | 4,830 (32.05%) |
| 40+ | 51 (3.07%) | 289 (2.60%) | 60 (2.86%) | 6 (2.93%) | 406 (2.69%) |
| Country |  |  |  |  |  |
| Nigeria | 93 (5.61%) | 886 (7.98%) | 330 (15.74%) | 17 (8.29%) | 1,326 (8.80%) |
| Pakistan | 1,159 (69.86%) | 8,170 (73.56%) | 1,530 (72.96%) | 166 (80.98%) | 11,025 (73.17%) |
| Tanzania | 297 (17.90%) | 1,557 (14.02%) | 161 (7.68%) | 14 (6.83%) | 2,029 (13.47%) |
| Zambia | 110 (6.63%) | 494 (4.45%) | 76 (3.62%) | 8 (3.90%) | 688 (4.57%) |
| Hemoglobin at baseline (g/L) (mean, SD) | 70.49 (13.88) | 83.67 (10.71) | 87.85 (9.05) | 79.27 (12.87) | 82.74 (11.84) |
| Anemia severity at baseline |  |  |  |  |  |
| Moderate | 895 (53.95%) | 9,931 (89.41%) | 1,999 (95.33%) | 164 (80.00%) | 12,989 (86.20%) |
| Severe | 764 (46.05%) | 1,176 (10.59%) | 98 (4.67%) | 41 (20.00%) | 2,079 (13.80%) |
| Multiple gestation | 88 (5.30%) | 402 (3.62%) | 84 (4.01%) | 9 (4.39%) | 583 (3.87%) |
| Gestational age (weeks) (mean, SD) | 36.84 (3.01) | 37.45 (2.62) | 37.52 (2.62) | 36.89 (3.14) | 37.39 (2.68) |
| Primiparity | 587 (35.40%) | 3,731 (33.59%) | 644 (30.71%) | 69 (33.82%) | 5,031 (33.39%) |
| Long-term infection | 83 (5.00%) | 385 (3.47%) | 74 (3.53%) | 10 (4.88%) | 552 (3.66%) |
| Placental abnormalities | 144 (8.68%) | 277 (2.49%) | 32 (1.53%) | 16 (7.80%) | 469 (3.11%) |
| Hypertensive disorder | 198 (11.93%) | 785 (7.07%) | 145 (6.91%) | 18 (8.78%) | 1,146 (7.61%) |
| Labor induction | 179 (10.79%) | 1,328 (11.96%) | 289 (13.78%) | 35 (17.07%) | 1,831 (12.15%) |
| Labor augmentation | 497 (29.96%) | 3,302 (29.73%) | 637 (30.38%) | 57 (27.80%) | 4,493 (29.82%) |
| Instrumental delivery | 78 (4.70%) | 302 (2.72%) | 38 (1.81%) | 4 (1.95%) | 422 (2.80%) |
| Episiotomy | 472 (28.45%) | 3,249 (29.25%) | 549 (26.18%) | 85 (41.46%) | 4,355 (28.90%) |
| Pharmacological pain relief | 148 (8.92%) | 1,411 (12.70%) | 247 (11.78%) | 4 (1.95%) | 1,810 (12.01%) |
| Duration of labor (hours) (median, IQR) | 8.83 (6.00-13.25) | 9.20 (6.33-13.75) | 9.64 (7.17-14.32) | 7.33 (5.08-11.62) | 9.18 (6.42-13.75) |
| Injury to the birth canal | 299 (18.02%) | 1,584 (14.26%) | 279 (13.30%) | 28 (13.66%) | 2,190 (14.53%) |
| Estimated blood loss (ml) (mean, SD) | 373.86 (307.51) | 302.41 (169.58) | 299.87 (161.41) | 331.51 (327.29) | 310.31 (192.72) |
| Clinically diagnosed PPH | 262 (15.79%) | 656 (5.91%) | 93 (4.43%) | 23 (11.33%) | 1,034 (6.86%) |
| Intervention for bleeding | 127 (7.66%) | 304 (2.74%) | 42 (2.00%) | 12 (5.91%) | 485 (3.22%) |
| Postpartum blood transfusion | 497 (29.96%) | 2,489 (22.41%) | 809 (38.58%) | 27 (13.17%) | 3,822 (25.37%) |
| Postpartum intravenous fluids | 689 (41.53%) | 3,320 (29.89%) | 658 (31.38%) | 97 (47.32%) | 4,764 (31.62%) |
| Baby admitted to NICU | 23 (1.39%) | 134 (1.21%) | 36 (1.72%) | 0 (0.00%) | 193 (1.28%) |
| Perinatal death before discharge | 260 (15.67%) | 758 (6.83%) | 125 (5.96%) | 29 (14.36%) | 1,172 (7.78%) |
| Small vulnerable newborn | 731 (44.17%) | 3,744 (33.78%) | 656 (31.34%) | 74 (36.27%) | 5,205 (34.62%) |
| Maternal death | 6 (0.36%) | 3 (0.03%) | 2 (0.10%) | 7 (3.45%) | 18 (0.12%) |
| Time to outcome assessment (hours) (median, IQR) | 20.48 (7.68-46.53) | 12.01 (6.19-23.48) | 10.49 (5.90-21.52) | 5.75 (2.48-9.03) | 12.34 (6.19-23.92) |
| SD – standard deviation; IQR – interquartile range; PPH – postpartum hemorrhage; NICU – neonatal intensive care unit. | | | | | |

Table S4. Distribution of maternal wellbeing outcome measures

| **Maternal wellbeing outcome** | **Median (IQR) (unless otherwise stated)** | **Prevalence ^1^**  **n / N (%)** |
| --- | --- | --- |
| Timed walk (meters) (mean, SD) | 212.17 (92.78) | - |
| Vigor (score) | 15 (10 to 19) | 11,402 / 15,068 (75.67%) |
| General fatigue (score) | 2 (0 to 4) | 1,321 / 15,068 (8.77%) |
| Physical fatigue (score) | 2 (0 to 5) | 1,364 / 15,068 (9.05%) |
| Mental fatigue (score) | 0 (0 to 0) | 417 / 15,068 (2.77%) |
| Emotional fatigue (score) | 0 (0 to 1) | 1,009 / 15,068 (6.70%) |
| Overall fatigue (score) | -9 (-16 to 1) | 3,952 / 15,068 (26.23%) |
| Other anemia symptoms (score) | 1 (0 to 3) | 1,113 / 15,068 (7.39%) |
| Difficulties with breastfeeding (score) | 0 (0 to 1) | 1,364 / 13,427 (9.89%) |
| Post-walk breathlessness (score) | 0 (0 to 1) | 129 / 14,488 (0.89%) |
| Expected difficulties with usual activities (score) | 0 (0 to 1) | 700 / 14,823 (4.72%) |
| Illness (score) | 0 (0 to 1) | 213 / 14,828 (1.44%) |
| Pain (score) | 0 (0 to 1) | 196 / 14,828 (1.32%) |
| The scores could range between 0 and 24 for the vigor and fatigue subscales, between -24 and 94 for the overall fatigue scale, between 0 and 28 for the other anemia symptoms scale, and between 0 and 4 for the remaining measures.  ^1^ Apart from overall fatigue, score measures were dichotomized according to whether women responded ‘quite a bit’ or ‘extremely’ to relevant Likert-style questions (table S1). For overall fatigue, ≥1 represents overall fatigue.  IQR – interquartile range; SD – standard deviation. | | |

Table S5. Unadjusted association between postpartum anemia and physical capacity, vigor, fatigue (general, physical, emotional, mental), other anemia symptoms, overall fatigue, difficulties breastfeeding, expected difficulties with usual activities, post-walk breathlessness, illness, and pain.

|  | **Postpartum anemia** | | |
| --- | --- | --- | --- |
|  | **Severe** | **Mild/ Moderate** | **None** |
|  | β-coefficient (95% CI) | |  |
| Physical capacity (six-minute walk) | -5.41 (8.77 to -2.04) | 0.00 | 6.64 (3.56 to 9.71) |
|  | Incident rate ratio (95% CI) | |  |
| Vigor | 0.93 (0.91 to 0.95) | 1.00 | 1.01 (0.99 to 1.03) |
| General fatigue | 1.32 (1.25 to 1.40) | 1.00 | 1.04 (0.98 to 1.10) |
| Physical fatigue | 1.27 (1.20 to 1.34) | 1.00 | 1.07 (1.02 to 1.12) |
| Emotional fatigue | 1.58 (1.39 to 1.80) | 1.00 | 1.17 (1.04 to 1.31) |
| Mental fatigue | 1.39 (1.22 to 1.59) | 1.00 | 1.21 (1.07 to 1.36) |
| Other anemia symptoms | 1.38 (1.30 to 1.46) | 1.00 | 1.04 (0.99 to 1.10) |
|  | Odds ratio (95% CI) | |  |
| Overall fatigue | 1.54 (1.35 to 1.77) | 1.00 | 1.12 (0.99 to 1.28) |
| Difficulties with breastfeeding | 1.16 (0.93 to 1.43) | 1.00 | 0.93 (0.78 to 1.11) |
| Expected difficulties with usual activities | 1.49 (1.18 to 1.88) | 1.00 | 0.97 (0.73 to 1.27) |
| Post-walk breathlessness | 2.93 (1.73 to 4.99) | 1.00 | 0.27 (0.06 to 1.13) |
| Illness | 1.37 (0.92 to 2.04) | 1.00 | 0.56 (0.34 to 0.92) |
| Pain | 0.96 (0.61 to 1.52) | 1.00 | 1.02 (0.65 to 1.60) |
| All models included a random effect for hospital. CI– confidence interval. | | | |

Table S6. Participants’ characteristics, by participation in the six-minute walk test

|  | **Six-minute walk test participation** | | |
| --- | --- | --- | --- |
|  | **Did not participate** | **Participated** | **Total** |
|  | **N=580** | **N=14,488** | **N=15,068** |
| Postpartum hemoglobin (g/L) (mean, SD) | 81.07 (14.59) | 85.92 (13.56) | 85.78 (13.61) |
| Postpartum anemia |  |  |  |
| No anemia (≥100 g/L) | 42 (10.10%) | 2,055 (14.22%) | 2,097 (14.11%) |
| Mild-Moderate (70-99 g/L) | 288 (69.23%) | 10,819 (74.89%) | 11,107 (74.73%) |
| Severe (<70 g/L) | 86 (20.67%) | 1,573 (10.89%) | 1,659 (11.16%) |
| Maternal age (years) | 27.48 (5.59) | 27.19 (5.61) | 27.20 (5.61) |
| Maternal age (years) |  |  |  |
| <20 | 25 (4.31%) | 870 (6.00%) | 895 (5.94%) |
| 20-29 | 340 (58.62%) | 8,597 (59.34%) | 8,937 (59.31%) |
| 30-39 | 198 (34.14%) | 4,632 (31.97%) | 4,830 (32.05%) |
| 40+ | 17 (2.93%) | 389 (2.68%) | 406 (2.69%) |
| Country |  |  |  |
| Nigeria | 40 (6.90%) | 1,286 (8.88%) | 1,326 (8.80%) |
| Pakistan | 504 (86.90%) | 10,521 (72.62%) | 11,025 (73.17%) |
| Tanzania | 21 (3.62%) | 2,008 (13.86%) | 2,029 (13.47%) |
| Zambia | 15 (2.59%) | 673 (4.65%) | 688 (4.57%) |
| Hemoglobin at baseline (g/L) | 77.77 (14.12) | 82.94 (11.70) | 82.74 (11.84) |
| Anemia severity at baseline |  |  |  |
| Moderate | 407 (70.17%) | 12,582 (86.84%) | 12,989 (86.20%) |
| Severe | 173 (29.83%) | 1,906 (13.16%) | 2,079 (13.80%) |
| Multiple gestation | 23 (3.97%) | 560 (3.87%) | 583 (3.87%) |
| Gestational age (weeks) (mean, SD) | 37.00 (2.84) | 37.40 (2.67) | 37.39 (2.68) |
| Primiparity | 228 (39.38%) | 4,803 (33.15%) | 5,031 (33.39%) |
| Long-term infection | 25 (4.31%) | 527 (3.64%) | 552 (3.66%) |
| Placental abnormalities | 45 (7.76%) | 424 (2.93%) | 469 (3.11%) |
| Hypertensive disorder | 71 (12.24%) | 1,075 (7.42%) | 1,146 (7.61%) |
| Labor induction | 120 (20.69%) | 1,711 (11.81%) | 1,831 (12.15%) |
| Labor augmentation | 253 (43.62%) | 4,240 (29.27%) | 4,493 (29.82%) |
| Instrumental delivery | 29 (5.00%) | 393 (2.71%) | 422 (2.80%) |
| Episiotomy | 192 (33.10%) | 4,163 (28.73%) | 4,355 (28.90%) |
| Pharmacological pain relief | 79 (13.62%) | 1,731 (11.95%) | 1,810 (12.01%) |
| Duration of labor (hours) (median, IQR) | 8.23 (5.00-13.42) | 9.23 (6.47-13.77) | 9.18 (6.42-13.75) |
| Injury to the birth canal | 74 (12.76%) | 2,116 (14.61%) | 2,190 (14.53%) |
| Estimated blood loss (ml) (mean, SD) | 364.26 (313.03) | 308.18 (186.11) | 310.31 (192.72) |
| Clinically diagnosed PPH | 91 (15.74%) | 943 (6.51%) | 1,034 (6.86%) |
| Intervention for PPH | 50 (8.65%) | 435 (3.00%) | 485 (3.22%) |
| Postpartum blood transfusion | 187 (32.24%) | 3,635 (25.09%) | 3,822 (25.37%) |
| Postpartum intravenous fluids | 294 (50.69%) | 10,010 (69.09%) | 10,304 (68.38%) |
| Baby admitted to NICU | 8 (1.38%) | 185 (1.28%) | 193 (1.28%) |
| Perinatal death before discharge | 90 (15.60%) | 1,082 (7.47%) | 1,172 (7.78%) |
| Small vulnerable newborn | 218 (37.72%) | 4,987 (34.49%) | 5,205 (34.62%) |
| Maternal death | 7 (1.21%) | 11 (0.08%) | 18 (0.12%) |
| Time to outcome assessment (hours) (median, IQR) | 7.76 (3.90-20.35) | 12.56 (6.23-23.99) | 12.34 (6.19-23.92) |
| SD – standard deviation; IQR – interquartile range; PPH – postpartum hemorrhage; NICU – neonatal intensive care unit. | | | |

Table S7. Unadjusted association between postpartum anemia and vigor, fatigue (general, physical, emotional, mental), and other anemia symptoms (sensitivity analysis using logistic regression to model count measures as binary outcomes)

|  | **Odds ratio (95% CI)** | | |
| --- | --- | --- | --- |
|  | **Postpartum anemia** | | |
| **Maternal wellbeing outcome** | **Severe** | **Mild/ Moderate** | **None** |
| Vigor | 0.68 (0.60 to 0.78) | 1.00 | 0.92 (0.82 to 1.04) |
| General fatigue | 1.69 (1.43 to 1.99) | 1.00 | 1.11 (0.93 to 1.33) |
| Physical fatigue | 1.65 (1.39 to 1.95) | 1.00 | 1.36 (1.14 to 1.63) |
| Emotional fatigue | 1.49 (1.24 to 1.81) | 1.00 | 1.04 (0.85 to 1.27) |
| Mental fatigue | 1.43 (1.06 to 1.93) | 1.00 | 1.47 (1.10 to 1.96) |
| Other anemia symptoms | 1.49 (1.24 to 1.78) | 1.00 | 1.06 (0.88 to 1.29) |
| All models included a random effect for hospital. CI – confidence interval. | | | |

Table S8. Adjusted association between postpartum anemia and vigor, fatigue (general, physical, emotional, mental), and other anemia symptoms (sensitivity analysis using logistic regression to model count measures as binary outcomes)

|  | **Odds ratio (95% CI)** | | |
| --- | --- | --- | --- |
|  | **Postpartum anemia** | | |
|  | **Severe** | **Mild/ Moderate** | **None** |
| Vigor | 0.89 (0.76 to 1.04) | 1.00 | 0.88 (0.77 to 1.00) |
| General fatigue | 1.39 (1.15 to 1.67) | 1.00 | 1.18 (0.98 to 1.42) |
| Physical fatigue | 1.45 (1.21 to 1.75) | 1.00 | 1.38 (1.15 to 1.66) |
| Emotional fatigue | 0.96 (0.76 to 1.22) | 1.00 | 1.16 (0.92 to 1.46) |
| Mental fatigue | 1.28 (0.92 to 1.77) | 1.00 | 1.48 (1.09 to 2.00) |
| Other anemia symptoms | 1.33 (1.09 to 1.62) | 1.00 | 1.10 (0.90 to 1.34) |
| All models included a random effect for hospital and adjust for time to outcome assessment, pre-birth anemia severity, age, gestational age, multiple gestation, primiparity, placental abnormalities, long-term infection, hypertensive disorder, labor induction, labor augmentation, pain relief, instrumental birth, episiotomy, trauma to birth canal, labor duration, estimated total blood loss (quadratic term), clinical diagnosis of postpartum hemorrhage, postpartum intravenous fluids, postpartum blood transfusion, small vulnerable newborn, and perinatal death before discharge. CI – confidence interval. | | | |

Table S9. Adjusted association between postpartum anemia and physical capacity, vigor, fatigue (general, physical, emotional, mental), other anemia symptoms, overall fatigue, difficulties breastfeeding, expected difficulties with usual activities, post-walk breathlessness, illness, and pain (sensitivity analysis that does not adjust for time at outcome assessment)

|  | **Postpartum anemia** | | |
| --- | --- | --- | --- |
|  | **Severe** | **Mild/Moderate** | **None** |
|  | β-coefficient (95% CI) | | |
| Physical capacity (six-minute walk) | -4.14 (-7.71 to -0.57) | 0.00 | 6.18 (3.08 to 9.27) |
|  | Incident rate ratio (95% CI) | | |
| Vigor | 0.99 (0.97 to 1.01) | 1.00 | 1.00 (0.99 to 1.02) |
| General fatigue | 1.19 (1.12 to 1.27) | 1.00 | 1.07 (1.01 to 1.13) |
| Physical fatigue | 1.19 (1.13 to 1.26) | 1.00 | 1.09 (1.03 to 1.14) |
| Emotional fatigue | 1.14 (1.01 to 1.29) | 1.00 | 1.23 (1.11 to 1.37) |
| Mental fatigue | 1.27 (1.10 to 1.46) | 1.00 | 1.22 (1.08 to 1.38) |
| Other anemia symptoms | 1.25 (1.18 to 1.33) | 1.00 | 1.07 (1.01 to 1.13) |
|  | Odds ratio (95% CI) | | |
| Overall fatigue | 1.20 (1.03 to 1.41) | 1.00 | 1.18 (1.03 to 1.35) |
| Difficulties with breastfeeding | 0.97 (0.77 to 1.24) | 1.00 | 1.00 (0.83 to 1.21) |
| Expected difficulties with usual activities | 1.55 (1.21 to 1.98) | 1.00 | 0.98 (0.75 to 1.29) |
| Post-walk breathlessness | 2.15 (1.22 to 3.77) | 1.00 | 0.47 (0.16 to 1.40) |
| Illness | 1.11 (0.74 to 1.67) | 1.00 | 0.66 (0.40 to 1.06) |
| Pain | 0.86 (0.54 to 1.38) | 1.00 | 1.19 (0.76 to 1.86) |
| All models included a random effect for hospital and adjust for pre-birth anemia severity, age, gestational age, multiple gestation, primiparity, placental abnormalities, long-term infection, hypertensive disorder, labor induction, labor augmentation, pain relief, instrumental birth, episiotomy, trauma to birth canal, labor duration, estimated total blood loss (quadratic term), clinical diagnosis of postpartum hemorrhage, postpartum intravenous fluids, postpartum blood transfusion, small vulnerable newborn, and perinatal death before discharge. CI – confidence interval. | | | |
